# Identical trends of SARS-Cov-2 transmission and retail and transit mobility during non-lockdown periods

**DOI:** 10.1101/2020.12.29.20248990

**Authors:** Bernard Cazelles, Catherine Comiskey, Benjamin Nguyen Van Yen, Clara Champagne, Benjamin Roche

## Abstract

Recent literature strongly supports the idea that mobility reduction and social distancing play a crucial role in transmission of SARS-Cov-2 infections. It was shown during the first wave that mobility restrictions reduce significantly infection transmission. Here we document the reverse relationship by showing, between the first two Covid-19 waves, a high positive correlation between the trends of SARS-Cov-2 transmission and mobility. These two trends oscillate simultaneously and increased mobility following lockdown relaxation has a significant positive relationship with increased transmission. From a public health perspective, these results highlight the importance of following the evolution of mobility when relaxing mitigation measures to anticipate the future evolution of the spread of the SARS-Cov-2.

---

Mobility of hosts and/or vectors has always had a large impact on the propagation of the transmission of diseases (Tizzoni et al., 2014; Findlater et al., 2018). This is once again the case for the Covid-19 pandemic (Kraemer et al., 2020).

Recent literature strongly supports the idea that mobility reduction and social distancing played a crucial role in the reduction of SARS-CoV-2 infections during the first wave. Some studies concluded that mobility patterns correlate with the prevalence of Covid-19 and that travel reduction had a positive effect on reducing the transmission of SARS-CoV-2 (Badr et al., 2020; Chang et al., 2020). Some other works have used the changes in mobility patterns to estimate the effect of mitigation measures on the reproductive number. They showed that the drop in mobility, overlapping with the introduction of mitigation measures induced a sharp reduction in transmission (Lemaitre et al., 2020; Park et al., 2020). But all these interesting results are limited to the first wave, before and during the lockdown period.

Nevertheless, it remains unknown if mobility increases following lockdown relaxation is actually associated with an enhanced transmission of SARS-Cov-2. Lockdowns impact a myriad of dimensions in our society, and not only on mobility patterns. This makes it difficult to draw a causal link between mobility and viral transmission from lockdown data. Instead, to better understand this link, we propose investigating this link between lockdown periods, to potentially use mobility data as an early signal for implementing public health measures.

**Here for the first time, to the best of our knowledge, we document a high correlation between the trends of SARS-Cov-2 transmission and mobility between the first two Covid-19 waves in some French regions and in Ireland**. We quantified the change in transmission of SARS-CoV-2 by computing the effective reproduction number *R*_*eff*_*(t)* with an original method based on a stochastic mechanistic model (Fig. A1) with time-varying parameters (Cazelles et al., 2018) and inferred using hospital data (Cazelles et al., 2020). The effective reproduction number is defined by the average number of secondary cases at time *t* arising from a primary infected case. It is an important metric for measuring time-varying transmissibility and assessing the effectiveness of different interventions. We used daily mobility data provided by Google, which is a proxy for real-time trends in movement patterns and human behavior (Google, 2020). These mobility data measure the percentage of change relative to the pre-pandemic baseline mobility and measure visits and the time spent in: retail and recreation, grocery and pharmacy, transit stations, workplace and residential.

Our analysis revealed that trends in transmission as estimated from well documented hospital data correlate strongly with trends in mobility patterns within and between the observed first and second epidemic waves for retail and recreation mobility, as well as transit station mobility. The *R*_*eff*_*(t)* increased and oscillated with mobility with a highly significant correlation between the 15^th^ of May and thee 15^th^ of October, with Pearson correlation coefficients above 0.5 (Figure). We show that these relationships are strong, particularly for retail and recreation mobility and transit station mobility. Additionally, we computed the Cross-Correlation Functions for these datasets that revealed the correlations were maximal for a delay between 0 and 10 days, and that this delay varied depending on the region (Figs A2-A3).

**Figure.**
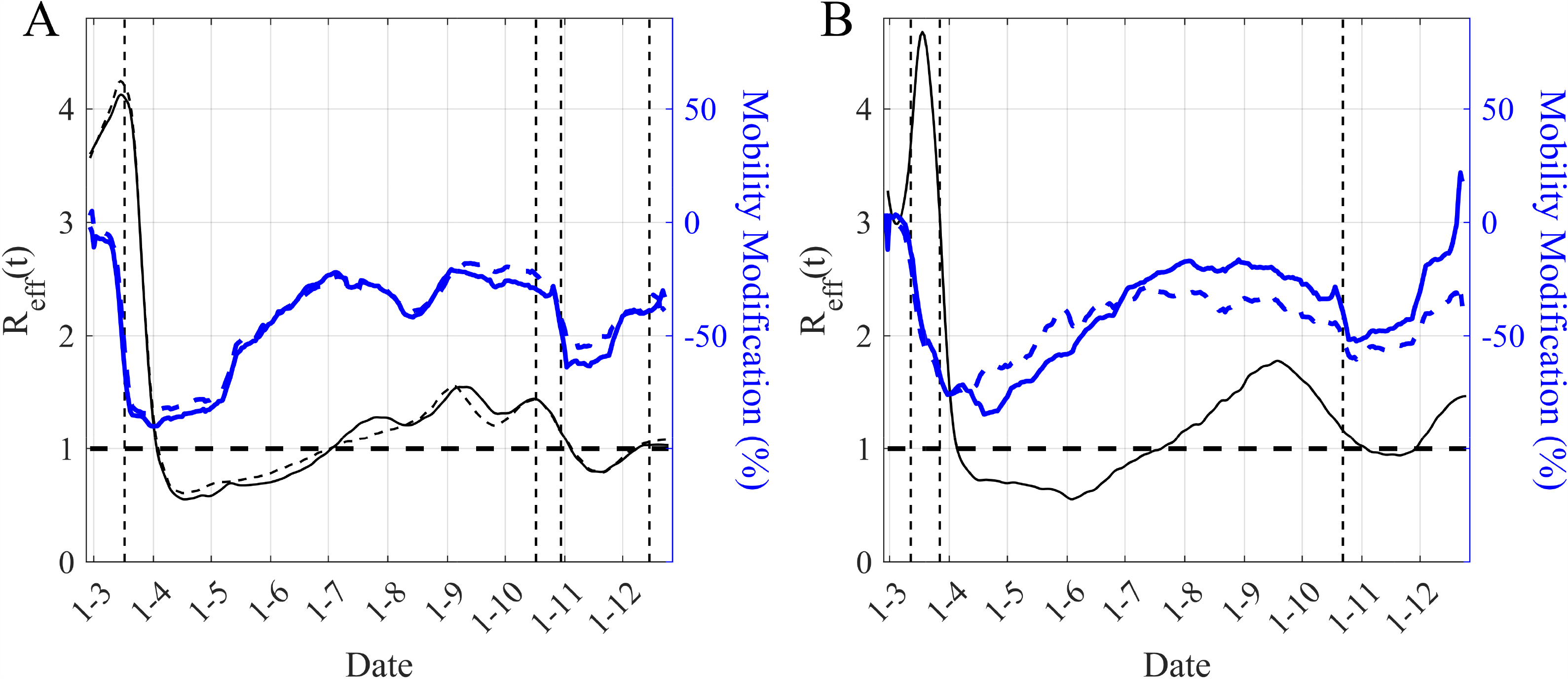
Time evolution of the estimated *R*_*eff*_*(t)* (black lines) and Google Mobility (retail and recreation mobility (continuous blue lines) and transport mobility (dashed blue lines)) in Ile-de-France region (A) and in Ireland (B). The mobility time series have been smoothed using moving average over a 7 days window. In A/ *R*_*eff*_*(t)* is computed for two different models with or without accounting for hospital discharges. Pearson Correlation in Ile-de-France (model with hospital discharges) with retail and recreation mobility 0.70 and with transport mobility 0.77. Pearson Correlation in Ile-de-France (model without hospital discharges) with retail and recreation mobility 0.64 and with transport mobility 0.70. Pearson Correlation in Ireland with retail and recreation mobility 0.86 and with transport mobility 0.56. Figs A2-A3 display the significance of the Cross-Correlation functions (CCF). Vertical dashed lines are main mitigation measures (lockdown and curfew) and the horizontal dashed lines are the threshold limit for *R*_*eff*_*(t)*.

Earlier findings showed that mobility reductions can dramatically reduce infections, our results now confirm the reverse relationship; increased mobility following lockdown relaxation has a significant, positive relationship with increased transmission. This suggests the importance of following the evolution of mobility data at different spatial resolutions to anticipate the future evolution of the spread of the SARS-Cov-2 and to guide public health policy.

Additionally, the strong relationship between retail and recreational mobility and increased transmissibility suggests that a return to baseline human activity poses a significant risk of increased infection. Moreover, even if a direct causal link cannot be drawn, these strong correlations indicate that an increase in retrial and recreational mobility promote high-intensity contact between people and also interaction between people of different households, resulting in increased SARS-CoV-2 transmission.

Our findings illustrate the importance of mobility at defined regional levels and perhaps more importantly, they highlight that mobility measures of transit and retail are more strongly correlated with transmission than other forms of mobility. These findings will aid the refinement of future possible lockdown policies and mitigation measures as we await the roll out of imminent vaccines.

## Data Availability

the data come from public data published online.

https://www.data.gouv.fr/fr/datasets/donnees-hospitalieres-relatives-a-lepidemie-de-covid-19/

https://covid19ireland-geohive.hub.arcgis.com/datasets/

https://www.google.com/covid19/mobility/

## Ethical approval

Not required.

---

**Figure A1.**
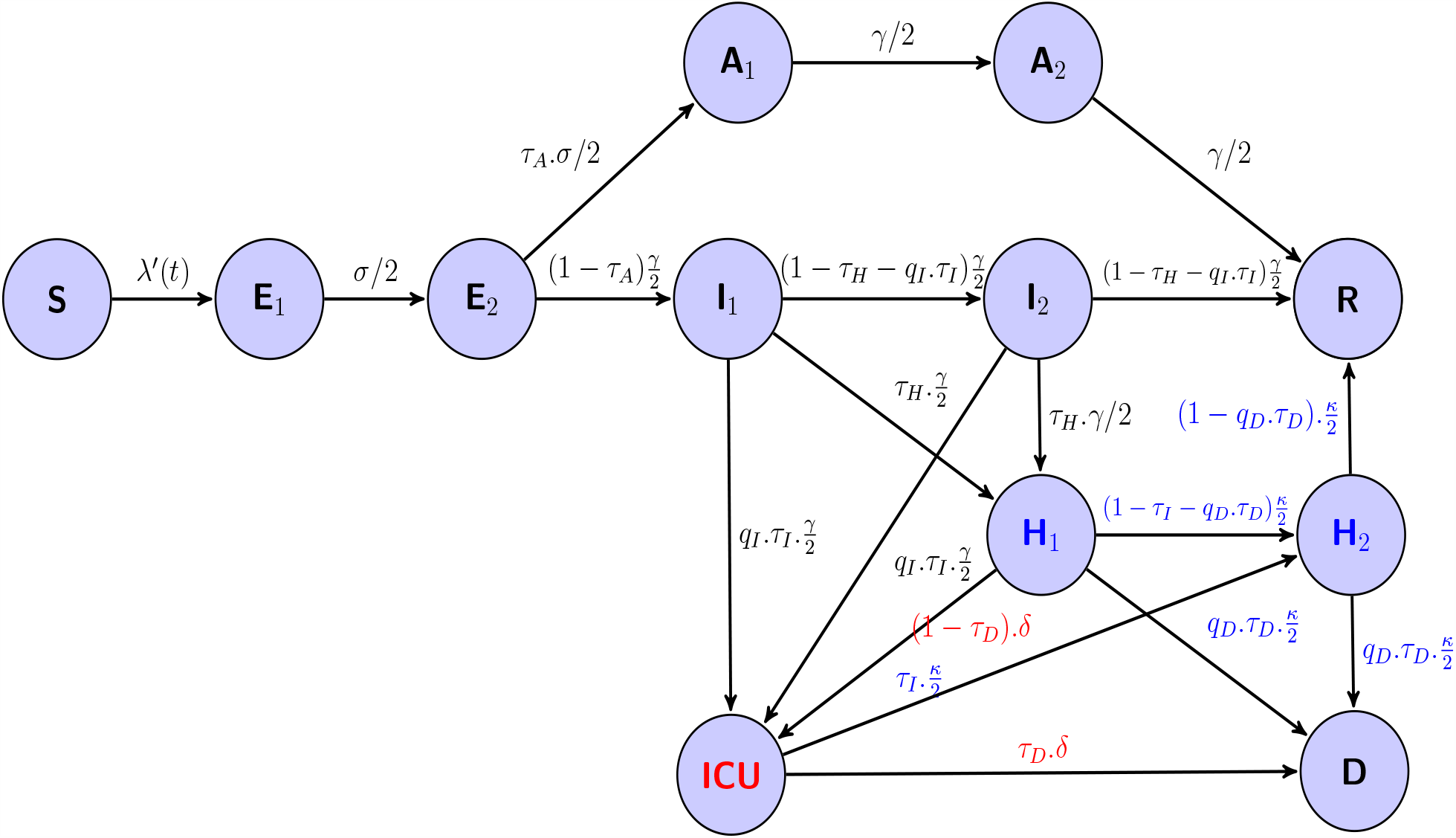
Flow diagram of the model used. It ncludes the following variables : the susceptibles S, the infected non-infectious E, the infectious symptomatic I, the infectious asymptomatic A, the removed people R, and the hospital variables : hospitalized people H, people in intensive care unit ICU, hospital discharge G, and deaths at hospital D. Erlang-distributed stage durations is also introduced for the E, I, A and H compartments to mimic a gamma distribution for stage duration in these compartments. *λ*′(*t*)= *β*(*t*).(*I*1+ q1.*I*2 + *q*2.(*A*1 + *A*2))/N then the force of infection is *λ*(*t*) = *λ′*(*t*).*S*(*t*). *β*(*t*) is the time-varying transmission rate, er the incubation rate, *γ* the recovery rate, 1/*κ* the average hospitalized period, 1/*δ* the average time spent in ICU, *τ*_*A*_ the fraction of asymptomatics, *τ*_*H*_ the fraction of infectious hospitalized, *τ*_*I*_ the fraction of ICU admission, *τ*_*D*_ the death rate, *q*1 and *q*2 the reduction of transmissibility of *I*2 and *A*_*i*_, *q*_*I*_ the reduction of the fraction of people admitted in ICU and *q*_*D*_ the reduction of the death rate. The subscripts 1 and 2 are for the 2 stages of the Erlang distribution of the considered variable. Flows in blue are from hospital (*H*_*i*_) and flow in red from *ICU*.

**Figure A2.**
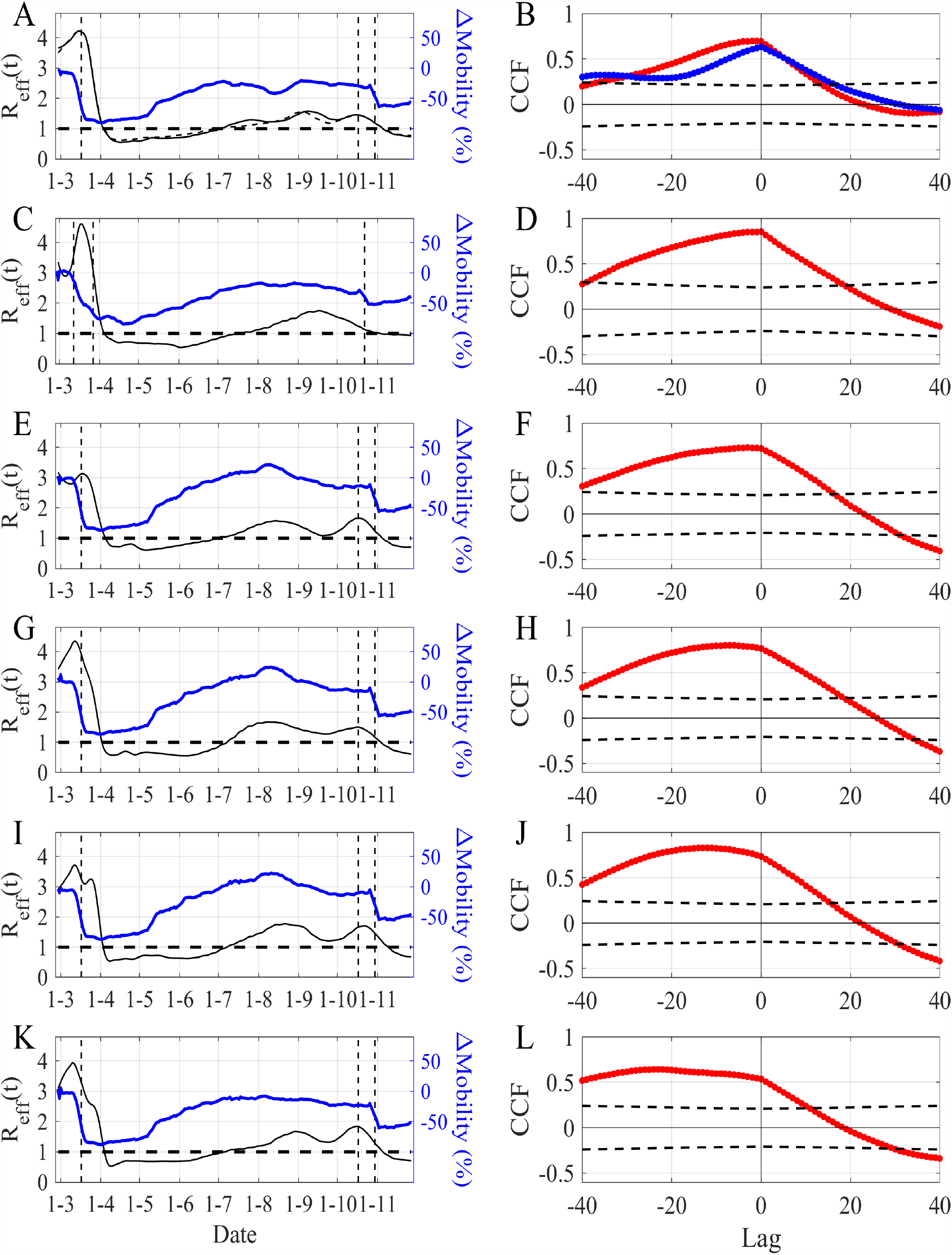
Correlations between the effective reproduction number and retail and recreation mobility. Left column : Time evolution of the estimated *R*_*eff*_ (*t*) (black line) and retail and recreation mobility (blue line). Right column : Cross-correlation functions between the estimated *R*_*eff*_ (*t*) and retail and recreation mobility computed between 15-05-2020 and 15-10-2020. The dashed black lines delimit the significant region at 0.1%. (A-B) Ile de France region, (C-D) Ireland ; (E-F) Provence Alpes Côte d’Azur region, (F-G) Occitanie region, (H-I) Nouvelle-Aquitaine region, (J-K) Auvergne Rhône Alpes region.

**Figure A3.**
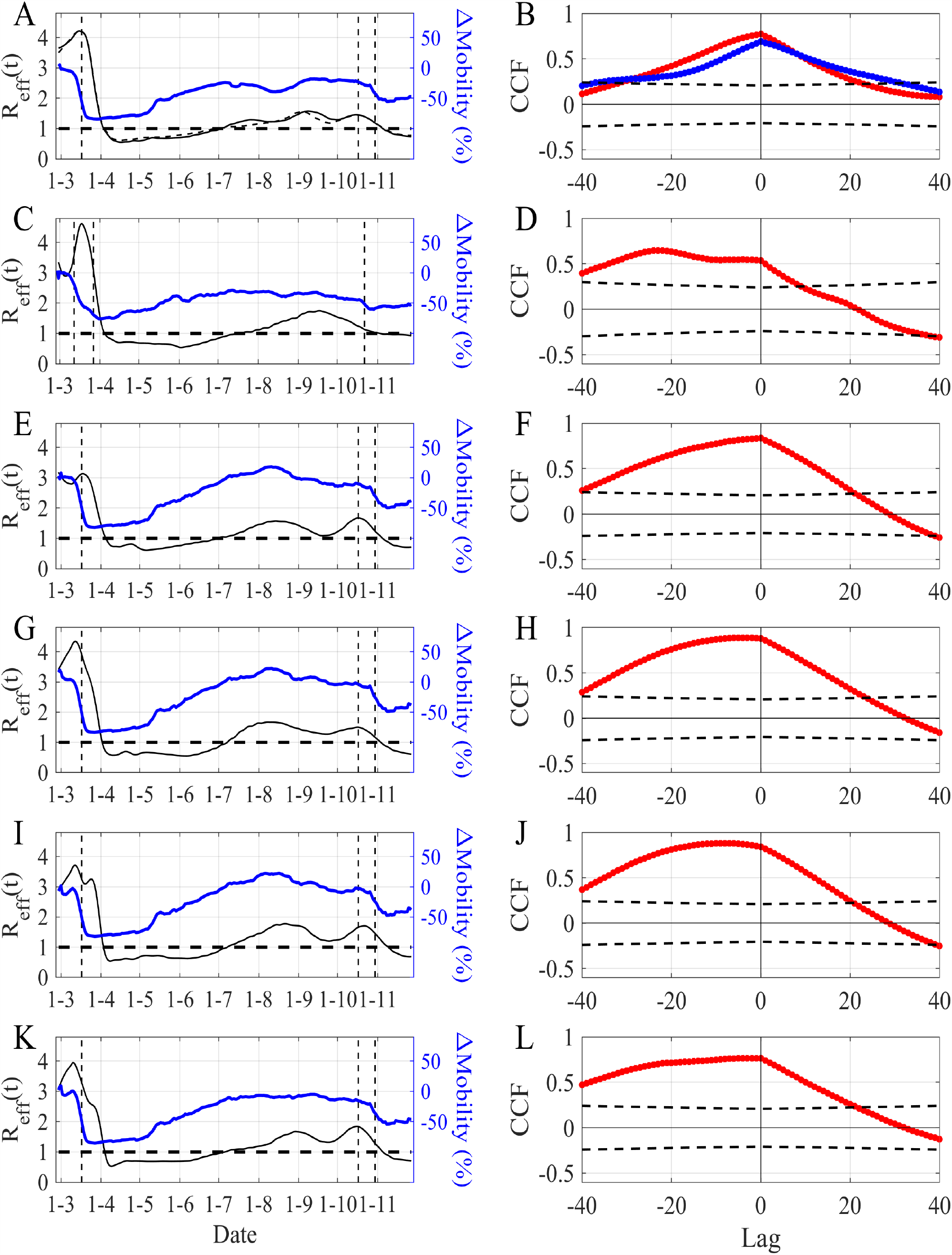
Correlations between the effective reproduction number and public transport mobility. Left column : Time evolution of the estimated *R*_*eff*_ (*t*) (black line) and public transport mobility (blue line). Right column : Cross-correlation functions between the estimated *R*_*eff*_ (*t*) and public transport mobility computed between 15-05-2020 and 15-10-2020. The dashed black lines delimit the significant region at 0.1%. (A-B) Ile de France region, (C-D) Ireland ; (E-F) Provence Alpes Côte d’Azur region, (F-G) Occitanie region, (H-I) Nouvelle-Aquitaine region, (J-K) Auvergne Rhône Alpes region.

